# What are the priority needs for those caring for children and young people with complex neurodisability within an ethnically diverse UK context? - The feasibility phase of the ENCOMPASS study

**DOI:** 10.1101/2023.03.23.23287248

**Authors:** Kirsten Prest, Emma Wilson, Io Vassiliadou, Sayeeda Ali, Monica Lakhanpaul, Christopher Morris, Cally Tann, Phillip Harniess, Sasha Lewis-Jackson, Hannah Kuper, Michelle Heys

**Author notes:** Correspondence, 2. UCL Great Ormond Street Institute of Child Health, University College London, 30 Guilford St, London WC1N 1EH, United Kingdom.

## Abstract

**Background:** Children and young people (CYP) with complex neurodisability experience multiple physical, communication, educational and social challenges which require complex packages of multidisciplinary care. Cerebral palsy, an exemplar complex neurodisability condition, is the most common cause of serious physical disabilities among children globally. It is unclear the best way to meet the emotional, social, practical, and empowerment needs of CYP with complex neurodisability and their caregivers. The aim of this study was to determine the needs and priorities of those caring for CYP with complex neurodisability. This forms part of the feasibility phase of a wider study (ENCOMPASS) which aims to adapt the ‘Baby Ubuntu’ intervention, a participatory caregiver programme for families of CYP with complex neurodisability, to an ethnically diverse urban UK context.

**Methods:** Two rounds of semi-structured interviews were conducted with 12 caregivers of CYP with complex neurodisability and six healthcare professionals from a variety of disciplines, recruited from a community child health service in London Borough of Newham, UK in 2020. The interviews included open-ended questions to explore caregiving trajectories, experiences of navigating health services and perceived service gaps. Transcripts were analysed using a data-driven inductive thematic analysis.

**Results:** Three themes were identified that related to the aim of understanding caregivers’ experiences and unmet needs relating to current service provision. These were (1) Caregiver Mental Health, (2) A Thirst for Knowledge and (3) The Need for Holistic Support. Mental health difficulties were reported, particularly around the period of diagnosis. Priority needs included the provision of clear information about the diagnosis and services offered, opportunities to forge peer support networks and for services across the community to collaborate.

**Conclusions:** The delivery of health services for CYP with neurodisability should encompass the broad needs of the family as well as meeting the clinical needs of the CYP.

## Background

Neurodisability refers to a group of long-term health conditions, either from birth or acquired, that describe functional limitations due to impairments of the brain and/ or the neuromuscular system (1). A neurodisability is said to be complex when the child or young person experiences physical/ motor impairments alongside cognitive, communication, educational and social needs which requires complex packages of multidisciplinary care (2). The diverse comorbidities that accompany such a diagnosis result in increased health services needs, particularly in those with more severe physical challenges (3).

Cerebral palsy, an exemplar complex neurodisability condition, is the most common cause of childhood physical disabilities globally, affecting 1.6 per 1000 live births in high-income countries and an estimated 3.4 per 1000 in low- and middle-income countries (4). Cerebral palsy is an umbrella term used when describing irreversible, non-progressive injuries to the foetal or infant’s brain. Children and young people with cerebral palsy present with multiple health, educational and social challenges (5) and resultant increased healthcare needs and utilisation (6)(7).

Healthcare professionals (HCPs) supporting children and young people with complex neurodisability are encouraged to work in partnership with caregivers (8). However, caregivers of children and young people with complex neurodisability also experience adverse health outcomes, for example, mental health and physical difficulties, which may result in higher levels of stress, anxiety, depression, pain, chronic diseases, financial instability, and reduced quality of life (9). Peer support is therefore also recommended for this caregiver population, which allows for opportunities to learn from others’ experiences, group problem-solve and discuss everyday issues (10)(11)(12). Multi-faceted family-centred interventions can positively influence the wellbeing of caregivers by providing practical and social support for the child or young person and their caregivers (13).

In the UK, guidelines from the National Institute of Health and Care Excellence (NICE) state that caregivers of children and young people with cerebral palsy require extensive support and up-to-date information about cerebral palsy and services, particularly around the time of diagnosis (5). However, the best way in which such support should be delivered to caregivers within the National Health Service (NHS) remains unclear.

Studies exploring caregivers’ experiences of accessing healthcare services for children and young people with complex disabilities indicate that the provision of emotional, social and informational support, or lack thereof, was as important as the medical aspect of care (14)(15)(16)(17). For example, when accessing rehabilitation services, parents of children and young people with cerebral palsy across low- and high-income contexts expressed a desire for information on understanding terms relating to cerebral palsy and how best to support their child’s development (18). Yet, caregivers frequently report difficulties in accessing information from health services about how to connect with other families (15)(16)(19)(20). Better coordination of health services is recommended for families of children with complex neurodisability to reduce parents’ experiences of repetitive support or lack of support (19).

Given the dearth of holistic, family-centred support for children and young people with complex neurodisability and their caregivers within the NHS (2), we reviewed interventions that might be suitable for this population in the UK (21)(22). From this review, we identified the <u>Ubuntu model</u> as a potential intervention that could be adapted to the UK context. “Ubuntu (previously: Getting to know Cerebral Palsy)” is a community-based participatory caregiver training programme that was adapted from a training resource called “Hambisela” in South Africa (23). “Ubuntu” has been developed and successfully tested and implemented in resource-limited settings. The modular, facilitated intervention is comprehensive in that it aims to promote inclusion and participation for the child with complex neurodisability in the community, to maximise the child’s health and development, to empower caregivers through information sharing and peer support, to address stigma, and to promote the human rights of children with disabilities (24). These holistic aims align with the WHO’s social determinants of health framework and the RAND “Future of Health” report in that social outcomes are considered as measures of improved health (25)(26). Evaluations of this programme in Ghana, Uganda, Brazil and Colombia demonstrated improvements in parental confidence and self-efficacy, as well as improved Quality of Life (QoL) for the child or young person and their families (27)(28)(29)(30).

The overall aim of this study was to describe the needs and priorities of those caring for children and young people with complex neurodisability in an ethnically diverse London borough and to identify gaps in current service provision.

### Setting

This was a UK-based study with caregivers accessing support for their children from HCPs in an NHS community setting. The NHS is a publicly funded healthcare service for residents of the UK and is free at the point of use. It is currently facing a workforce crisis, which has worsened throughout the COVID-19 pandemic (31). This study was based in the ethnically diverse borough of Newham in East London. In the UK, ethnic minorities often experience unequal access to hospital services and poorer patient experience compared to majority white populations (32)(33). Newham has the lowest proportion of first-language English speakers across England and Wales (34) and an estimated 52% of children live in poverty, higher than any other London borough (35). Adults in Newham experience a significantly higher prevalence of poor mental and physical health, as well as higher service use compared to the rest of the UK (36).

## Methods

This study is the first step of three in assessing the feasibility of co-adapting the “Baby Ubuntu” programme to the UK. It forms part of the feasibility phase of the MRC complex intervention development framework (37) and the first step in the ADAPT framework for adapting interventions to other contexts (39). Steps two and three (future publications) will describe theoretical acceptability and feasibility and suggestions on how to adapt content and delivery. Ethical approval was obtained from the Health Research Authority (ref 20/YH/0311).

This was a qualitative study, framed by a social constructionism paradigm which assumes that human experiences cannot be understood without the knowledge of their societal context, which includes language, culture, and power relations (38). We note the impact of ethnicity, language, culture, stigma and views on disability in relation to the experiences of the caregiver participants in this study. We conducted semi-structured interviews in two rounds between January and April 2021 with caregivers of children with complex neurodisability and healthcare professionals (HCPs). We provided detailed information about the “Ubuntu” program at the start of the interviews, and participants could ask questions for clarification. The interview guides had open-ended questions about caregiving trajectories, navigating health services, and service gaps.

Caregivers were recruited using research and clinical databases, as well as advertising and social media. Participants were eligible if they were the adult caregiver of a child or young person with a diagnosed complex neurodisability, aged 18 years or younger and resided in Newham, East London.

Using a purposive sampling approach, we identified 45 caregivers of children and young people with complex neurodisability from clinical lists who were contacted by a clinician. Twenty agreed to be contacted by a researcher and eight agreed to participate in the study. A second round of recruitment through parent/carer forums and contacting non-responders resulted in a further eight participants. In total, 16 caregivers consented to participate and 12 attended at least one interview. Reasons for refusal included limited time, moving out of borough and competing childcare needs.

To increase ethnic and linguistic diversity of the recruitment of participants, it was made clear that translators could be available if English was not their first language. Translating services were used for the initial clinician recruitment calls, however there were a few instances where a translator was not available.

We aimed to recruit healthcare professionals from a range of disciplines. We invited seven HCPs providing care to children and young people with complex neurodisability in Newham. Of these, six agreed to participate.

All participants provided informed consent via an online form. One female researcher (IV) conducted all interviews remotely via Microsoft Teams due to COVID-19 restrictions (39). The researcher is young (29 years), Greek Cypriot and a trained clinical psychologist. The interviews were video-recorded and transcribed verbatim. Some participants requested the support of family members to assist with translation. A short questionnaire was used to collect demographic information before the interview.

Data analysis was managed using NVivo software. We conducted a data-driven inductive thematic analysis (40). IV and SLJ read all transcripts (n=18) to familiarise themselves with the data before coding them to create an initial coding framework, from which emergent themes and sub-themes were generated. These transcripts were then coded by a third researcher (KP), and the coding framework was further developed and refined. MH, EW and KP met to discuss coding labels and definitions. An additional meeting was held with EW and KP to discuss the final themes, sub-themes, analytical conclusions and to come to a consensus if there were any disagreements (41).

Patient and Public Involvement was incorporated in the formative stages of the study. Local clinicians consulted families of children with complex neurodisability to identify unmet needs and gaps in service provision, which informed the study protocol and grant application. A workshop was held for patients and the public to provide feedback on patient-facing materials.

## Results

### Participant characteristics

Semi-structured interviews were conducted with six HCPs and 12 caregivers. Demographic information was collected (See Tables 1, 2 and 3). Caregivers’ ages ranged from 31-42 years, with their children’s ages ranging from 2-15 years. Most of the caregivers were mothers from ethnic minority groups, with variations in types of accommodation, employment and state benefits received. Two participants did not complete the demographic questionnaire. Demographic data was collected for the CYP whose caregivers consented to share their information (n=8).

**Table 1:** Participant Demographic Information: Healthcare Professionals.

| **Healthcare Professional Disciplines** | ***n*=6** |
| --- | --- |
| Health Visitor | 2 |
| Occupational Therapist | 1 |
| Paediatrician | 1 |
| Physiotherapist | 1 |
| Speech and Language Therapist | 1 |

**Table 2:** Participant Demographic Information: Parents and Carers.

| **Demographic information of parents and carers** | ***n*=12** |
| --- | --- |
| *Caregiver's age, range (mean)* | 31 – 42 (38) years |
| *Gender* |  |
| Female | 10 |
| Male | 2 |
| *Marital status* |  |
| Single, never married | 3 |
| Married, or in a domestic partnership | 7 |
| Unknown | 2 |
| *Ethnic group* |  |
| Any White Background | 4 |
| Afro-Caribbean | 2 |
| Pakistani | 3 |
| Bangladeshi | 1 |
| Unknown | 2 |
| *Type of accommodation* |  |
| Flat/ apartment/ maisonette | 5 |
| House/ bungalow | 5 |
| Unknown | 2 |
| *Own or rent accommodation* |  |
| Rents | 6 |
| Owns | 4 |
| Unknown | 2 |
| *Number of biological children living with caregivers, range (mean)* | 1-4 (2.4) |
| *Highest Qualification* |  |
| GCSE/ O-Level equivalent | 4 |
| Bachelor's degree | 5 |
| Master's degree | 1 |
| Unknown | 2 |
| *Employment* |  |
| Part-time paid employment | 2 |
| Full-time paid employment | 3 |
| Unpaid or voluntary work including looking after family | 5 |
| Unknown | 2 |
| *State benefits received* |  |
| Disability Living Allowance | 9 |
| Motability Scheme | 2 |
| Carer's Allowance | 5 |
| Child Support | 1 |
| Universal Credit | 5 |
| Unknown | 2 |

**Table 3:** Participant Demographic Information: Children and Young People. \**GMFCS: Gross Motor Function Classification System records a child’s functional level of movement* (42). These data were only available for five of the children and young people.

|  |  |
| --- | --- |
| **Demographic information of children and young people** | ***n*=8** |
| *Age, range (mean)* | 2 – 15 (8) years |
| *Gender* |  |
| Female | 1 |
| Male | 7 |
| *Diagnosis* |  |
| Cerebral Palsy | 7 |
| Other neurodisability | 1 |
| *GMFCS\** |  |
| I | 1 |
| II | 1 |
| III | 1 |
| IV | 1 |
| V | 1 |
| Unknown | 3 |

### Themes

Three themes were identified that related to the aim of understanding caregivers’ experiences and unmet needs. These were (1) Caregiver Mental Health, (2) A Thirst for Knowledge and (3) The Need for Holistic Support.

#### Caregiver Mental Health

Several factors were described by caregivers and HCPs as adversely impacting upon caregiver mental health, namely: receipt of diagnosis; the overwhelming number of tasks required of them; feeding; concern about the future; transition to school; and feelings of isolation. Conversely adequate support, knowledge and early diagnosis were reported by two participants as positively impacting on their mental health.

Many caregivers described feelings of anxiety and low mood, particularly around the period of diagnosis. These reported difficulties with mental health were irrespective of the severity of their child’s diagnosis or the caregiver characteristics.

> *“I think being a parent is overwhelming…but I think when you add this into the mix when you weren’t expecting it, because most of us don’t expect it. It is overwhelming. I’ve definitely had periods of feeling low and feeling like a failure and then wanting to give up. -C13*

All participants reported their own or their partner’s initial denial over their child’s diagnosis. This experience further influenced their mental health, often resulting in periods of anxiety and depression.

> *“I did have a bit of a small breakdown after about a month when my husband accepted what had happened to my son. I think I let my guard down knowing that and then it kind of hit me all in one go. I had a little bit of a panic attack once*.*” -C16*

They reported feeling powerless within the process and overwhelmed by the many tasks required of them. Caregivers described the burden of appointments for their child, including difficulty keeping track of them all and the overloading of information. A few mentioned that they had no time for themselves.

Multiple caregivers discussed feeding issues as a particular trigger for anxiety as part of the overall pressure felt in caring for their child. One parent reported feeling worried about leaving the house as they did not know whether places would be accessible for their child.

Worrying about the future was another aspect of anxiety that caregivers frequently mentioned. One caregiver reported feeling powerless as they did not know what to expect and what their child might be able to do. Other caregivers expressed worry about their child’s plans for schooling and becoming adults.

> *I’m worried a lot about college…Now everything is okay, but when he finishes school and he has to go to college and start living his adult life, I’m really worried and I don’t*
>
> *know how everything will be. It’s basically something new for me and it worries me a lot. -C11*

One HCP confirmed the above by reporting their observations of caregivers feeling anxious about sending their child to school and transitioning between different education environments.

> *From working with families, [the education side of things] is always quite anxiety provoking. The whole way through transitioning into different education systems and making sure that the child’s at the right school and [receiving] the right opportunities in school. And I think that’s quite hard for some families to get their head around. -H4*

As part of the impact on caregivers’ mental health, many described how their friends and family had difficulty understanding their situations. Caregivers reported struggling to socialise with other families because of their child’s barriers to play. They discussed having no prior reference point to how to interact with a disabled child, or not knowing what to say to the parent/caregiver.

> *In my social group I am the only one with a child with special needs. So even when I try to speak to some friends, they’ll never understand. And they always look at the brighter side of everything, because he’s always so happy. But then they don’t see when we have to do physio every day, when I’m struggling feeding him. They don’t see the dark side of it. They just see the happy-go-lucky baby. -C3*

The varying difficulties reported above resulted in parents feeling isolated and that their support networks could not understand them. This sentiment of isolation was shared amongst many of the participants, which was reported to be compounded by the lack of support and information provided by HCPs.

> *Isolation is such a big theme amongst parents and carers with kids with disabilities*.*-C1*

One caregiver reported a markedly different experience. They described themselves and their partner as the ‘luckiest parents’ as they received an early diagnosis and increased support and advice from doctors and allied health professionals. They reported that this increased support assisted them in their acceptance of the diagnosis. Another parent described how their anxiety was reduced when HCPs provided them with information about what to expect for certain milestones and activities. -C11.

> *I think that anxiety and depression is such a major thing amongst carer parents that if we are given enough information to help us just calm down, you would find that the rates of depression anxiety would lower a little bit as well. -C1*

#### A Thirst for Knowledge

A prominent finding was that caregivers expressed a lack of knowledge about their child’s condition, associated co-morbidities and the accompanying services available, along with frustration with the overuse of medical jargon by HCPs. They described seeking information and assurance from sources outside of healthcare.

Caregivers had varying experiences of how involved HCPs were in the initial stages of their journey, but many reported feeling disappointed by the system.

> *I felt a bit let down by the medical system. I found out that my son probably has cerebral palsy before I was told because the doctors didn’t give me any information…I felt let down by NHS staff when I initially got the diagnosis. I was very angry and frustrated. In the beginning, the first NHS staff that kind of made me feel better was my physiotherapist a few months later. – C16*

Caregivers reported difficulties understanding what the diagnosis of cerebral palsy would mean for their child, along with accompanying co-morbidities such as feeding and communication difficulties, bowel problems or epilepsy.

> *We were sort of thrown the word cerebral palsy, there wasn’t ever an explanation. I wasn’t sent any information packs*… *I was [in my 20s] at the time, so I had no idea what was going on and you can feel really lost. -C1*

A few caregivers described various supports available to families, however, there was often a lack of knowledge about how to access them. This included a lack of awareness of assistive devices and equipment availability, financial support such as carers’ allowance, and support groups. From the caregivers’ perspective, it was sometimes difficult to understand the different roles of the various HCPs.

> *There is support out there, but not a lot of people know where to look for the support, because I didn’t, and I still don’t know. -C3*

One HCP commented that information about the local offer for families could be challenging to find.

> *I think the local offer is not good in Newham compared to other areas and it’s hard to find information in it. I struggled to find information myself when I was looking for things. - H1*

One of the most frequently mentioned difficulties was the complex terminology that caregivers had to dissect after their child was diagnosed with cerebral palsy. This included medical jargon and acronyms, such as the commonly used Gross Motor Functioning Classification System (GMFCS, (42)) levels. Language barriers made this particularly challenging. Caregivers expressed concern that there was not enough time during doctors’ appointments to deal with all of their questions, particularly in the period just after diagnosis. One parent reportedly felt more relaxed if they could bring someone else to the appointment.

> *I just went on to some parent forums because lots of the NHS staff I found to be more medical. And there was lots of jargon that I didn’t understand and that depresses me a bit because I’m not a stupid person. I then just joined forums and started talking to parents as I just didn’t know what else to do. -C13*

Due to the gaps in knowledge described above, caregivers frequently conducted independent research. Sources of information included the internet, charities, and parent groups. Concerns were raised about processing the volume of information available. Caregivers reported feeling fearful of their child’s prognosis when seeing children with severe difficulties online.

> *I had no idea what was going on and you can feel really lost because you’re told that they have a condition and then the problem is when you go on the Internet, there is such a vast world [of information]…So I’m seeing cerebral palsy people [who are not]*
>
> *walking, in wheelchairs and not feeding…when you are a new parent or even a parent whose never had a child [with a] disability before, I feel as though you need to be given proper information so that you’re not running away with it yourself. -C1*
>
> *Some parents will actively tell you that they’re looking on Google and others will say that they’re not ready to do that -H2*

#### The Need for Holistic Support

All participants described the need to provide holistic and timely support for families with a child with complex neurodisability – both from the healthcare system and from the community.

Participants discussed the lack of communication between various departments who care for children and young people with complex neurodisability. It was suggested that collaborative joint-working of HCPs in varying disciplines should be prioritised to streamline services for families. Caregivers described the need to seek support elsewhere, for example privately or in different countries. HCPs described the tension of a service under pressure, against a desire to do more to support families. Multiple caregivers raised concerns about the infrequency of appointments, and it was suggested that having a telephone number to call and speak to someone, would be welcome.

> *I was walking into [that first appointment] on my own. I have my little boy with me and when I left, I just sobbed because I had absolutely no idea that it was coming. And there was no support. There was nothing. The child’s got cerebral palsy. Not even a telephone number of a support group. Just absolute darkness. And I was very low for a long while until I dragged myself up because I didn’t know what to do. -C13*

All participants raised the need for individualised support for children with complex neurodisability, particularly concerning feeding, positioning, home visits and condition-specific information. One HCP described how services were often aimed at different diagnoses and that there was a clear need for further individualised support for families with children and young people with cerebral palsy.

> *A lot of these coaching parental support things are around, but for autism and behavioural issues, not for children with cerebral palsy, so that’s what’s missing. Their needs are very, very different and that needs to be addressed in a different way-H4*

Caregivers stated the need for further support from the community, and to connect with others who have lived experience. Most caregivers reported that it was important to hear stories and learn from others in similar situations, even though each family’s needs are unique. This connection and learning reportedly reduced feelings of isolation and positively influenced their mental health.

> *I think the group delivery is quite important because, especially at the beginning, you feel like you are the only person who is going through it, so it’s nice to hear when there’s other people coming in with similar backgrounds and stories. Part of this whole thing is you can feel really isolated so even something as simple as doing a group exercise, I think psychologically will do something. -C1*

A few caregivers expressed a desire to help other families who were earlier on in their journeys, to assist in alleviating worries. One participant mentioned the importance of having adult role models who have a diagnosis of cerebral palsy. It was suggested that involving families or people with cerebral palsy in the training workshops would be valuable.

> *When I first heard the word cerebral palsy, I didn’t know what it was, I didn’t know anything about it, it was so new to me. And thankfully when I read a story, exactly what I’m going be producing as well, it helped me to understand. -C5*

Along with the support required from health services and groups, a few caregivers expressed the need for further support from the wider community and schools. Both caregivers and HCPs described the benefits of extending educational support and awareness to friends and family members, along with the wider community, with the overall aim of decreasing stigma and improving understanding.

> *I think it would also be helpful [to invite] your extended family [to the groups] … it’s sometimes important for your circle of family and friends to understand what it is that you’re going through on a day-to-day basis. -C14*
>
> *Once people understand what disability is all about, maybe what cerebral palsy is all about, they don’t go around judging and looking. -H6*

Multiple caregivers expressed a desire for support and advice from healthcare services about the schooling process and how to access funding. Parents/caregivers discussed the need for Special Educational Needs Co-ordinators (SENCOs) to have specific training, to improve their understanding of the needs of children and young people with complex neurodisability. It was suggested that schools in the area, both specialist and mainstream, could be linked into the programme. Finally, caregivers described the need for more leisure opportunities for their children, for example sports clubs, to improve overall participation in the community.

## Discussion

This qualitative study explored needs and priorities for those caring for children and young people with neurodisability in a diverse urban setting. To our knowledge, it is one of a handful of studies exploring these factors in such a diverse population. We report significant unmet needs, resulting in subsequent adverse impacts on caregiver mental health, consistent with studies in upper-middle and high-income contexts (9)(8)(43)(44)(45). Recent research in the field of child neurodisability emphasises the importance of supporting families as a whole (46). Our data suggest a community-based intervention, such as the “Ubuntu” model, may meet some of these unmet needs and provide family-centred support.

We define unmet needs in healthcare to broadly cover non-use, delayed use and sub-optimal use of health services, which can have detrimental impacts to the wellbeing of certain populations (47). Caregivers’ unmet needs in this study included the need for support for their mental health, the need to be empowered with knowledge and skills, and the need for services to be better co-ordinated to enhance the overall care. Healthcare professionals identified gaps in service provision for individualised support for children and young people with cerebral palsy and their families.

One of the ways in which caregivers’ mental health was impacted by having a child or young person with a complex neurodisability, was through the difficulties in managing multiple appointments from a variety of healthcare services. McCann et al. (48) conducted a systematic review of the daily patterns of time use for children with complex needs and found that caregivers spent considerable time engaged in healthcare-related activities for their children, placing a significant caregiving burden on them. With relation to mental health, the concept of ‘future’ was mentioned frequently in this study as caregivers expressed anxiety about the unknown potential of their child’s development. Rosenbaum and Gorter (49) place an important emphasis on HCPs keeping a child’s future in mind when working with families, without ignoring present concerns. An implication for practice might be for HCPs to incorporate discussions around future into consultations, as this might have an impact on caregiver mental health.

Previous studies conducted in upper-middle and high-income contexts have reported a link between poor mental health, social isolation, exclusion, and stigma and the time-consuming, continuous tasks required of caregivers (50)(51)(52)(45)(53). Caregivers in this study reported feeling isolated and alone in their difficulties, with their support networks not able to understand their struggles. This experience is mirrored in a variety of contexts and demonstrates the importance of forging support networks for caregivers. It should not be assumed that a caregivers’ ability to cope is correlated with the nature or severity of their child or young person’s diagnosis, which is supported by previous studies (54). Caregivers in this study expressed a need to connect with others in similar situations. Not only does this provide opportunities for a shared social identify which can have a positive impact on health and wellbeing and bring a sense of belonging (55), but it also provides opportunities for mutual support. Reciprocity offered in social support was important to participants, which is echoed in previous literature (10) (56).

NICE guidelines recommend the provision of timely and appropriate information for families of children with cerebral palsy, particularly during the period around diagnosis (5). Multiple caregivers in this study reported the need for further information and individualised support about their child’s condition. This is echoed by previous studies, particularly in relation to parents’ finding it challenging to receive up-to-date information and support (19)(43)(57). It is also recommended that the support provided for families of children with cerebral palsy be individualised, as a ‘one-size-fits-all’ approach is not appropriate for this population (8)(58). Caregivers further described the need for support in understanding complex medical terminology. Parents from a variety of contexts have raised similar concerns (18)(20) emphasising the need for clear communication of medical terms, particularly considering language and culture (43).

A key finding of this study was the need to improve co-ordination of services in healthcare, schools and in communities, particularly during significant transition points. A review into the quality of care provided for patients with chronic neurodisability in the UK (2) emphasised the critical importance of improving communication between HCPs and families, as well as streamlining the co-ordination of care amongst the varying multi-disciplinary teams. Previous studies have also emphasised how crucial it is to improve co-ordination of the varying services that aim to meet the multi-faceted needs of these families (19).

### Strengths and Limitations

This study had several strengths, such as involving both service users and healthcare providers, allowing for multiple perspectives to be gained. Despite a small sample size, there was variation in caregiver and child characteristics, leading to rich data generation. The interviewer’s (IV) nationality, particularly being an immigrant, allowed rapport to be built quickly, while her clinical psychology training helped her discuss sensitive topics. Some interviews were conducted with family members acting as interpreters, which could lead to a loss of nuance and privacy, however allowing participants to answer in their first language is a strength. Some caregivers were not provided with study information due to a lack of interpreters during recruitment, meaning data from underrepresented families may be missing. Finally, only one child did not have a diagnosis of cerebral palsy, raising questions about combining children with different neurodisability conditions in a group.

The COVID-19 pandemic affected recruitment and data collection, with remote collection being more convenient for some, but challenging for others due to privacy or digital access. Three participants didn’t attend online interviews, and those who refused to participate cited time and childcare pressures, possibly excluding those with the greatest caregiving burden.

Findings of this study may be transferable and relevant to other ethnically diverse urban settings, with similar pressures on NHS health and social service provision.

## Conclusion

This study demonstrated the importance of focusing on caregivers’ mental health, empowering them with knowledge, and streamlining services within the healthcare setting and the wider community to better support families of children and young people with neurodisability. The delivery of health services should encompass the broad needs of the family, by providing holistic care with clear and up-to-date information, facilitating the forging of peer support networks, as well as meeting the biomedical needs of the children and young people. Some of these priorities can be addressed through community-based caregiver programmes.

## Data Availability

All data produced in the present study are available upon reasonable request to the authors

## References

(1) Morris C, Janssens A, Tomlinson R, Williams J, Logan S. Towards a definition of neurodisability: a Delphi survey. Dev Med Child Neurol 2013 Dec;55(12):1103-1108.

(2) The National Confidential Enquiry into Patient Outcome and Death. Each and Every Need: A review of the quality of care provided to patients aged 0-25 years old with chronic neurodisability, using the cerebral palsies as examples of chronic neurodisabling conditions. 2018; Available at: https://www.ncepod.org.uk/2018cn.html. Accessed 14 March 2023.

(3) Carter B, Bennett CV, Jones H, Bethel J, Perra O, Wang T, et al. Healthcare use by children and young adults with cerebral palsy. Developmental medicine and child neurology 2021 Jan;63(1):75–80.

(4) McIntyre S, Goldsmith S, Webb A, Ehlinger V, Hollung SJ, McConnell K, et al. Global prevalence of cerebral palsy: A systematic analysis. Dev Med Child Neurol 2022 Dec;64(12):1494-1506.

(5) National Institute for Health and Care Excellence. Cerebral palsy in under 25s: assessment and management. [NG62]. 2017 May 17,;31(38):15.

(6) Meehan E, Williams K, Reid SM, Freed GL, Babl FE, Sewell JR, et al. Comparing emergency department presentations among children with cerebral palsy with general childhood presentations: a data linkage study. Dev Med Child Neurol 2017 -08-08;59(11):p1188.

(7) Berry JG, Poduri A, Bonkowsky JL, Zhou J, Graham DA, Welch C, et al. Trends in Resource Utilization by Children with Neurological Impairment in the United States Inpatient Health Care System: A Repeat Cross-Sectional Study. PLoS Medicine 2012 Jan;9(1):e1001158.

(8) Hayles E, Harvey D, Plummer D, Jones A. Parents’ Experiences of Health Care for Their Children With Cerebral Palsy. Qual Health Res 2015 Aug;25(8):1139-1154.

(9) Pousada M, Guillamón N, Hernández-Encuentra E, Muñoz E, Redolar D, Boixadós M, et al. Impact of Caring for a Child with Cerebral Palsy on the Quality of Life of Parents: A Systematic Review of the Literature. J Dev Phys Disabil 2013 Oct;25(5):545–577.

(10) Shilling V, Morris C, Thompson-Coon J, Ukoumunne O, Rogers M, Logan S. Peer support for parents of children with chronic disabling conditions: a systematic review of quantitative and qualitative studies. Dev Med Child Neurol 2013;55(7):602–609.

(11) Law M, King S, Stewart D, King G. The Perceived Effects of Parent-Led Support Groups for Parents of Children with Disabilities. Phys Occup Ther Pediatr 2002;21(2-3):29–48.

(12) Kingsnorth S, Gall C, Beayni S, Rigby P. Parents as transition experts? Qualitative findings from a pilot parent-led peer support group. Child: Care, Health and Development 2011 Nov;37(6):833–840.

(13) Prest KR, Borek AJ, Boylan AR. Play-based groups for children with cerebral palsy and their parents: A qualitative interview study about the impact on mothers’ well-being. Child: care, health and development. 2022; 48(4), 578 –587.https://doi.org/10.1111/cch.12962

(14) Campbell D, Milne J, Garg P, Tomsic G, Ong N, Silove N. Parents’ and Service Providers’ Experiences of Accessing Health Services from an Intellectual Disability Health Team. J Prim Care Community Health 2021;12:21501327211014068.

(15) Anderson M, Elliott EJ, Zurynski YA. Australian families living with rare disease: experiences of diagnosis, health services use and needs for psychosocial support. Orphanet Journal of Rare Diseases 2013 Feb 11,;8(1):22.

(16) Bellin MH, Osteen P, Heffernan C, Levy JM, Snyder-Vogel ME. Parent and Health Care Professional Perspectives on Family-centered Care for Children with Special Health Care Needs: Are We on the Same Page? Health & social work 2011 Nov;36(4):281–290.

(17) Lindsay S, King G, Klassen AF, Esses V, Stachel M. Working with immigrant families raising a child with a disability: challenges and recommendations for healthcare and community service providers. Disability and rehabilitation 2012 Nov;34(23):2007-2017.

(18) Jindal P, Macdermid JC, Rosenbaum P, Direzze B, Narayan A. Perspectives on rehabilitation of children with cerebral palsy: exploring a cross-cultural view of parents from India and Canada using the international classification of functioning, disability and health. Disability and Rehabilitation 2017 -07-26;40(23):2745.

(19) Kiernan G, Courtney E, Ryan K, McQuillan R, Guerin S. Parents’ experiences of services for their child with a life-limiting neurodevelopmental disability. Children’s health care 2020 Apr 02,;49(2):134–152.

(20) Liptak GS, Orlando M, Yingling JT, Theurer-Kaufman KL, Malay DP, Tompkins LA, et al. Satisfaction With Primary Health Care Received by Families of Children With Developmental Disabilities. Journal of pediatric health care 2006;20(4):245–252.

(21) Heys M, Lakhanpaul M, Allaham S, Manikam L, Owugha J, Oulton K, et al. Community-based family and carer-support programmes for children with disabilities. Paediatrics and Child Health 2020;30(5):180–185.

(22) Wilson E, Kuper H, Tann CJ, Heys M et al. Rapid Response: Reverse innovation in practice: the Getting to Know Cerebral Palsy programme. BMJ. 2019;367.

(23) van Aswegen T, Myezwa H, Potterton J, Stewart A. The effect of the Hambisela programme on stress levels and quality of life of primary caregivers of children with cerebral palsy: A pilot study. S Afr J Physiother 2019 Feb 20;75(1):461.

(24) Tann CJ, Kohli-Lynch M, Nalugya R, Sadoo S, Martin K, Lassman R, et al. Surviving and Thriving: Early Intervention for Neonatal Survivors With Developmental Disability in Uganda. Infants Young Child 2021 Jan;34(1):17–32.

(25) Corbett J, d’Angelo C, Gangitano L, Freeman J. Future of Health: Findings from a survey of stakeholders on the future of health and healthcare in England. Rand Health Q 2018 Apr 1;7(3):1.

(26) World Health Organisation. A conceptual framework for action on the social determinants of health. 2010. Available from: https://apps.who.int/iris/handle/10665/44489. Accessed 14 March 2023.

(27) Sadoo S, Nalugya R, Lassman R, Kohli-Lynch M, Chariot G, Davies HG, et al. Early detection and intervention for young children with early developmental disabilities in Western Uganda: a mixed-methods evaluation. BMC Pediatrics 2022;22(1):158.

(28) Zuurmond M, David O’Banion, Gladstone M, Carsamar S, Kerac M, Baltussen M, et al. Evaluating the impact of a community-based parent training programme for children with cerebral palsy in Ghana. PLoS ONE 2018;13(9):e0202096.

(29) Duttine A, Smythe T, Calheiros de Sá MR, Ferrite S, Moreira ME, Kuper H. Assessment of the feasibility of Juntos: A support programme for families of children affected by Congenital Zika Syndrome. Wellcome Open Research 2022;7(77).

(30) Smythe T, Reichenberger V, Pinzón EM, Hurtado IC, Rubiano L, Kuper H. The feasibility of establishing parent support groups for children with congenital Zika syndrome and their families: a mixed-methods study. Wellcome Open Research 2021;6(158).

(31) The King’s Fund. The NHS in a nutshell. 2022; Available at: https://www.kingsfund.org.uk/projects/nhs-in-a-nutshell. Accessed 23 November, 2022.

(32) Nazroo JY, Falaschetti E, Pierce M, Primatesta P. Ethnic inequalities in access to and outcomes of healthcare: analysis of the Health Survey for England. J Epidemiol Community Health 2009 Dec;63(12):1022-1027.

(33) Raleigh V HJ. The health of people from ethnic minority groups in England | The King’s Fund. 2021; Available at: https://www.kingsfund.org.uk/publications/health-people-ethnic-minority-groups-england Accessed 14 March 2023.

(34) Aston-Mansfield. Newham: Key Statistics. A detailed profile of key statistics about Newham by Aston-Mansfield’s Community Involvement Unit. 2017; Available at: https://www.aston-mansfield.org.uk/wp-content/themes/aston_mansfield/uploads/Newham_Statistics_.pdf2017. Accessed 14 March 2023.

(35) Trust for London. Poverty and inequality data for Newham. 2020; Available at: https://www.trustforlondon.org.uk/data/boroughs/newham-poverty-and-inequality-indicators/. Accessed 14 March 2023.

(36) Pevalin DJ. Socio-economic inequalities in health and service utilization in the London Borough of Newham. Public Health 2007 Aug;121(8):596–602.

(37) Skivington K, Matthews L, Simpson SA, Craig P, Baird J, Blazeby JM, et al. Framework for the development and evaluation of complex interventions: gap analysis, workshop and consultation-informed update. Health Technol Assess 2021 Sep;25(57):1–132.

(38) Burr V. Social Constructionism. 3rd Edition ed. London: Routledge; 2015.

(39) Manikam L, Allaham S, Zakieh O, Bou Karim Y, Demel I, Ali S, et al. Online community engagement in response to COVID-19 pandemic. Health Expect 2021;24(2):728–730.

(40) Braun V, Clarke V. Using thematic analysis in psychology. Qualitative Research in Psychology 2006 Dec 1,;3(2):77–101.

(41) Boyatzis RE. Transforming qualitative information: Thematic analysis and code development. Thousand Oaks, CA, US: Sage Publications, Inc; 1998.

(42) Palisano R, Rosenbaum P, Walter S, Russell D, Wood E, Galuppi B. Development and reliability of a system to classify gross motor function in children with cerebral palsy. Developmental Medicine & Child Neurology 1997 Apr;39(4):214–223.

(43) Arfa S, Solvang K, Berg B, Jahnsen R. Disabled and immigrant, a double minority challenge: a qualitative study about the experiences of immigrant parents of children with disabilities navigating health and rehabilitation services in Norway. BMC Health Serv Res. 2020; 20(134). https://doi.org/10.1186/s12913-020-5004-2

(44) Gilson K, Johnson S, Davis E, Brunton S, Swift E, Reddihough D, et al. Supporting the mental health of mothers of children with a disability: Health professional perceptions of need, role, and challenges. Child Care Health Dev 2018;44(5):721–729.

(45) Rodrigues SA, Fontanella BJB, Avó LRS, Germano CMR, Melo DG. A qualitative study about quality of life in Brazilian families with children who have severe or profound intellectual disability. J Appl Res Intellect Disabil 2019;32(2):413–426.

(46) Rosenbaum PL, Novak-Pavlic M. Parenting a Child with a Neurodevelopmental Disorder. Curr Dev Disord Rep 2021;8(4):212–218.

(47) Smith S, Connolly S. Re-thinking unmet need for health care: introducing a dynamic perspective. Health economics, policy and law 2020 Oct;15(4):440–457.

(48) McCann D, Bull R, Winzenberg T. The daily patterns of time use for parents of children with complex needs: A systematic review. J Child Health Care 2012;16(1):26–52.

(49) Rosenbaum P, Gorter JW. The ‘F-words’ in childhood disability: I swear this is how we should think. Child: Care, Health and Development 2012 Jul;38(4):457–463.

(50) Resch JA, Mireles G, Benz MR, Grenwelge C, Peterson R, Zhang D. Giving Parents a Voice. Rehabilitation Psychology 2010 May;55(2):139–150.

(51) Breitkreuz R, Wunderli L, Savage A, Mcconnell D. Rethinking resilience in families of children with disabilities: a socioecological approach. Community, Work & Family 2014;17(3):1–20.

(52) Khanlou N, Mustafa N, Vazquez LM, Davidson D, Yoshida K. Mothering children with developmental disabilities: A critical perspective on health promotion. Health Care Women Int 2017;38(6):613–634.

(53) Luijkx J, Putten AAJ, Vlaskamp C. Time use of parents raising children with severe or profound intellectual and multiple disabilities. Child Care Health Dev 2017;43(4):518–526.

(54) Parkes J, Caravale B, Marcelli M, Franco F, Colver A. Parenting stress and children with cerebral palsy: a European cross-sectional survey. Developmental Medicine & Child Neurology 2011 Sep;53(9):815–821.

(55) Haslam SA, Jetten J, Postmes T, Haslam C. Social Identity, Health and Well-Being: An Emerging Agenda for Applied Psychology. Applied psychology 2009 Jan;58(1):1–23.

(56) Reid A, Imrie H, Brouwer E, Clutton S, Evans J, Russell D, et al. “If I Knew Then What I Know Now”: Parents’ Reflections on Raising a Child with Cerebral Palsy. Phys Occup Ther Pediatr 2011;31(2):169–183.

(57) © Care Quality Commission. Special Review: Health care for disabled children and young people. 2012 March. Available from:https://www.cqc.org.uk/sites/default/files/documents/health_care_for_disabled_children.pdf Accessed 14 March 2023.

(58) Terwiel M, Alsem MW, Siebes RC, Bieleman K, Verhoef M, Ketelaar M. Family-centred service: differences in what parents of children with cerebral palsy rate important. Child: care, health and development 2017;43(5):663–669.

